# Personalized qEEG-guided tDCS/tRNS in children with autism spectrum disorder: an exploratory randomized sham-controlled trial (NCT02004236)

**DOI:** 10.64898/2026.11.26.26369099

**Authors:** Moises Aguilar Domingo, Joaquín Pérez Navarro, Hector Alfaro Madrid, Citlalli Mellado Balanzar, Principal Investigator — Brainmech Foundation

**Affiliations:** Spanish Foundation for Neurometrics Development (Valencia, Spain, 46021); Moises Aguilar Domingo (Liverpool, United Kingdom, L4 5QL); BrainMech Foundation; Bigdata4brain LTD

## Abstract

**Introduction:** Autism Spectrum Disorder (ASD) presents marked neurobiological and clinical heterogeneity that limits the efficacy of standardized interventions. Non-invasive transcranial electrical neuromodulation (tDCS/tRNS) guided by quantitative electroencephalography (qEEG) and event-related potentials (ERPs) may allow personalization of the intervention on language and social cognition networks. Given the absence of prior individualized qEEG-guided tDCS/tRNS trials in pediatric ASD, this study is reported as an exploratory proof-of-mechanism trial; clinical efficacy estimates require independent replication.

**Methods:** Interventional, prospective, randomized, double-blind, sham-controlled exploratory clinical trial with three parallel arms (N = 225 recruited; 209 completed; 100% male; approximately 75% right-handed and 25% left-handed or functionally ambidextrous). Three arms are compared: 1) Experimental: tRNS/tDCS fronto-temporal cortex (n=75), 2) Experimental: tRNS/tDCS over fusiform temporal cortex / individualized qEEG montage (n=75), and 3) Placebo Comparator: sham (n=75 initiated; n=59 completed), evaluating longitudinal stability at weeks 24 and 48. This study is reported in accordance with CONSORT guidelines for randomized trials, with stimulation parameters described per TIDieR recommendations.

**Results:** The principal findings of this exploratory trial are: (1) pediatric safety — no serious adverse events (0/209) and only transient, self-limited dizziness in 1.33% and 2.67% of active arms versus 0% in sham; and (2) target engagement — sLORETA showed significant reduction of excess Theta current density (t = −6.84 to −7.12, p < 0.0001) and maturation of Alpha rhythm in Wernicke’s area (T5), accompanied by normalization of interhemispheric and focal coherence. Clinical between-group comparisons showed large but exploratory signals on CARS/EVAI (d = 4.92–5.15, p < 0.0001), which require independent replication. Notably, the individualized qEEG-guided montage did not improve D-KEFS verbal fluency relative to sham (10.5 ± 1.6 → 10.4 ± 1.1 vs. 9.9 ± 1.1 → 10.0 ± 0.9; between-group difference not significant), and the fixed frontotemporal montage showed a decline (9.3 ± 1.3 → 8.3 ± 1.2). The cognitive signal is therefore null/ambiguous, and baseline D-KEFS imbalance limits interpretation of cognitive outcomes. N170 latency shortened from ∼210–215 ms to 188–190 ms (pooled d = 1.52, p < 0.0001) and amplitude increased from ∼1.4 µV to ∼2.9–3.1 µV (d = 1.48–1.62), but these ERP findings should be considered hypothesis-generating.

**Conclusions:** qEEG-guided transcranial neuromodulation in pediatric ASD showed a favorable safety profile and evidence of target engagement on sLORETA and coherence measures. Clinical efficacy signals were large but exploratory and require independent replication in adequately powered, fully blinded trials. The absence of a cognitive benefit for the individualized montage and the baseline imbalance in D-KEFS scores underscore the need for cautious interpretation.

## 1. Introduction, Demographic Characterization, and Empirical Evidence

Autism Spectrum Disorder (ASD) is a neurodevelopmental condition characterized by persistent deficits in communication and social interaction, along with restricted and repetitive behavior patterns. In accordance with the protocol registered in <u>ClinicalTrials.gov</u> NCT02004236 (tDCS25112013) [1,2], the present approach targets a pediatric population aged 5 to 12 years who meet DSM-IV diagnostic criteria for ASD [3] (with at least 2 years of prior diagnostic evolution) and without associated intellectual disability or neurological pathology.

**Given the absence of prior individualized qEEG-guided tDCS/tRNS trials in pediatric ASD, this study was designed and is reported as an exploratory proof-of-mechanism trial.** Clinical efficacy estimates derived from this trial require independent replication, and the primary contributions of this report are pediatric safety data and evidence of target engagement using qEEG, sLORETA, and coherence mapping.

### 1.1 Demographic Characterization and Laterality of the Cohort (NCT02004236)

In the NCT02004236 clinical trial [2], 225 pediatric participants were recruited (75 per arm), with 209 subjects completing the protocol (100% retention in active arms and 78.67% in the sham arm). The demographic and baseline neuropsychological characterization of the cohort presents the following parameters:

- **Sex and Age Distribution:** 100% male (209/209 in the analysed sample in Spain), with a global mean age of 7.2 ± 2.12 years (7.5 ± 1.98 years in the frontotemporal arm; 6.9 ± 2.1 years in the fusiform temporal arm; and 7.2 ± 2.3 years in the sham arm), stratified into three age bands: 4–6 years (mean 5.06 ± 0.91 years), 7–9 years (mean 7.8 ± 0.91 years), and 10–12 years (mean 10.7 ± 0.68 years).
- **Hand Dominance (Hemispheric Lateralization):** The laterality distribution of the clinical sample shows approximately **75% right-handed and 25% left-handed or functionally ambidextrous** participants (crossed laterality included; the 1% discrepancy in the original report reflects rounding). This is a critical parameter that determines the individualized anodic–cathodic polarity (anode at T5 to cathode at T4 in right-handed patients; anode at T6 to cathode at T3 in left-handed patients).

### 1.2. Clinical, Spectral, Effect Size, and Follow-up Results (Weeks 24 vs. 48)

The comparative analysis between active neuromodulation arms and the sham group shows convergence between behavioral measures (CARS/EVAI scales [5,6] and D-KEFS verbal fluency [7]), spectral reorganization of qEEG [8,9], functional coherence, and cognitive evoked potentials (P3b wave and N170/N196-P236 complex) [10,11]. However, **these between-group differences are exploratory and should not be interpreted as confirmatory efficacy estimates.**

#### Baseline imbalance

Baseline D-KEFS scores differed across arms (Fusiform/qEEG 10.5 ± 1.6 vs. Frontotemporal 9.3 ± 1.3 vs. Sham 9.9 ± 1.1). Post-hoc adjustment for baseline scores attenuated the between-group cognitive effect. The primary signal in this trial is therefore electrophysiological and clinical, not cognitive.

#### D-KEFS result

Contrary to the expected cognitive benefit, the individualized qEEG-guided montage **did not improve D-KEFS verbal fluency relative to sham** (10.5 ± 1.6 → 10.4 ± 1.1 vs. 9.9 ± 1.1 → 10.0 ± 0.9; between-group difference not significant). The fixed frontotemporal montage showed a decline (9.3 ± 1.3 → 8.3 ± 1.2). The previously reported d = 2.18 for D-KEFS at weeks 24–48 likely reflects inflated between-group variance rather than a true cognitive effect, and should be interpreted with caution given the baseline imbalance. **The cognitive signal is therefore null/ambiguous.**

### 1.3. Longitudinal Evolution Week 24 vs. Week 48 and Electrophysiological Correlation

Table 2 details the persistence of clinical and neurophysiological effects in the extended follow-up (Week 24 vs. Week 48) comparing individualized active neuromodulation versus the placebo/sham arm, along with effect size (Cohen’s d) and electrophysiological correlation (Pearson’s r) with respect to symptomatic reduction in CARS/EVAI [5,6]. **These longitudinal findings are exploratory and hypothesis-generating.**

**Table 1.** Clinical, Spectral, and Evoked Potential Biomarkers by Arm (NCT02004236) [. **2,5–11]**

| **Parameter / Biomarker** | **tRNS/tDCS Fronto-temporal (n=75)** | **tRNS/tDCS Fusiform / qEEG (n=75)** | **Sham (n=59\*)** | **Effect Size (d) / Correlation** |
|---|---|---|---|---|
| **Sex / Cohort Laterality** | 100% Male \| 75% Right / 26% Left | 100% Male \| 75% Right / 26% Left | 100% Male \| 75% Right / 26% Left | Homogeneous (p > 0.05) |
| **Global Mean Age (SD)** | 7.5 (1.98) | 6.9 (2.10) | 7.2 (2.30) | Global: 7.2 (2.12) |
| **Finalization Rate / Dropouts** | 75/75 (100%) / 0% | 75/75 (100%) / 0% | 59/75 (78.67%) / 16 (21.33%) | Chi2 = 34.2; p < 0.0001 |
| **CARS / EVAI (Pre to Post)** | 30.6 (1.9) to 19.4 (2.1) | 32.1 (2.2) to 20.2 (1.8) | 31.8 (3.4) to 30.1 (2.5) | d = 4.92 - 5.15 (p < 0.0001) |
| **D-KEFS Verbal Fluency (Pre to Post)** | 9.3 (1.3) to 8.3 (1.2) | 10.5 (1.6) to 10.4 (1.1) | 9.9 (1.1) to 10.0 (0.9) | d = 0.38 (Fixed montage) |
| **Theta Power T5 (uV, Pre to Post)** | 3.20 (0.30) to 2.20 (0.41) | 2.70 (0.11) to 1.50 (0.36) | 3.50 (0.50) to 3.30 (0.84) | d = 1.98 - 3.24 \| r = 0.71 |
| **Posterior Alpha Power (uV)** | 1.15 (0.22) to 2.35 (0.31) | 1.10 (0.19) to 2.52 (0.28) | 1.18 (0.21) to 1.22 (0.24) | d = 4.05 - 4.98 \| r = -0.68 |
| **Beta Power (uV)** | 0.13 (0.03) to 0.18 (0.04) | 0.09 (0.02) to 0.11 (0.03) | 0.13 (0.03) to 0.13 (0.03) | Spectral normalization |
| **Monastra Theta/Beta Ratio** | 25.0 (0.88) to 12.0 (1.10) | 30.0 (1.10) to 14.0 (0.52) | 26.0 (0.53) to 25.0 (0.77) | d = 13.85 - 15.20 \| r = 0.79 |
| **ECPT Reaction Time (ms)** | 756 (23.5) to 690 (10.4) | 698 (34.2) to 556 (7.4) | 722 (18.9) to 714 (3.2) | d = 3.12 - 9.85 (p < 0.0001) |
| **ECPT Omission / VCPT (Post)** | 85 (5.2) / 68 (2.3) | 91 (3.2) / 57 (4.7) | 88 (4.6) / 20 (1.8)** | p < 0.001 |
| **VCPT Commission (Pre / Post)** | 6 (0.2) / 6 (0.2) | 8 (1.1) / 8 (1.1) | 19 (0.9) / 19 (0.9) | d = 15.1 - 18.4 (p < 0.0001) |
| **P3b Amplitude (uV)** | 0.50 (0.12) to 2.10 (0.25) | 0.70 (0.40) to 1.80 (0.65) | 0.30 (0.20) to 0.40 (0.32) | d = 4.18 - 5.64 \| r = -0.66 |
| **N170 / N196-P236 (uV)** | 1.45 (0.35) to 2.85 (0.42) | 1.38 (0.31) to 3.12 (0.48) | 1.42 (0.33) to 1.48 (0.36) | d = 1.48 - 1.62 \| r = -0.74 |
| **SAEs / Dizziness (1-2 h)** | 0/75 (0%) / 1/75 (1.33%) | 0/75 (0%) / 2/75 (2.67%) | 0/59 (0%) / 0/59 (0%) | No SAEs (0/209) |

Note: p < 0.0001 (mixed ANOVA / Bonferroni-adjusted between-group t-test). Data derived from NCT02004236 (N=225; n=209 completed) [2]. † Effect sizes for these exploratory outcomes are reported for transparency but should not be interpreted as confirmatory efficacy estimates; they require independent replication. ‡ Values are absolute amplitudes (μV). N170 is a negative-going component; negative peak amplitudes are reported in the text (e.g., −1.4 to −2.8 μV). \*\* p < 0.001.

**Table 2.** Longitudinal Follow-up at Weeks 24 and 48 [. **2,8,9].**

| **Variable** | **Active Week 24** | **Active Week 48** | **Sham Week 24** | **Sham Week 48** | **Effect Size (d) Week 48** | **Pre-Post r** |
|---|---|---|---|---|---|---|
| **CARS / EVAI (Total Score)** | 19.2 +/- 2.0 | 19.8 +/- 2.2 (95.2%) | 30.4 +/- 2.6 | 30.9 +/- 2.8 | d = 4.51 (p < 0.0001) | Clinical reference |
| **D-KEFS Verbal Fluency** | 14.8 +/- 1.9 | 14.5 +/- 2.0 | 10.1 +/- 1.2 | 10.0 +/- 1.3 | d = 2.18 (p < 0.0001) | r = -0.69 (p < 0.001) |
| **Theta / Beta Ratio** | 11.8 +/- 1.0 | 12.3 +/- 1.2 | 25.2 +/- 0.8 | 25.6 +/- 0.9 | d = 12.45 (p < 0.0001) | r = +0.79 (p < 0.0001) |
| **Relative Posterior Alpha (%)** | 38.4 +/- 4.2% | 37.6 +/- 4.5% | 19.2 +/- 3.1% | 18.9 +/- 3.3% | d = 4.72 (p < 0.0001) | r = -0.68 (p < 0.001) |
| **Slow Focal Coherence (Z)** | +0.42 +/- 0.31 | +0.48 +/- 0.35 | +2.35 +/- 0.44 | +2.41 +/- 0.46 | d = 1.45 (p < 0.0001) | r = +0.64 (p < 0.001) |
| **Interhemispheric Alpha (Z)** | -0.28 +/- 0.29 | -0.34 +/- 0.32 | -2.18 +/- 0.41 | -2.24 +/- 0.43 | d = 1.38 (p < 0.0001) | r = -0.71 (p < 0.0001) |
| **N170 / N196-P236 (Latency ms)** | 188 +/- 11 ms | 190 +/- 12 ms | 226 +/- 15 ms | 228 +/- 16 ms | d = 1.52 (p < 0.0001) | r = -0.74 (p < 0.0001) |

Note: Negative t values (blue) indicate significant reduction of hyperactivity (cortical noise); positive values (green) indicate maturation of Alpha rhythm. \*\* p < 0.001, \*\*\* p < 0.001. † Exploratory effect estimates; require independent replication.

**Table 3.** Intervention Arms.

| **Arm** | **Arm Type** | **Intervention / Device** | **N Recruited / Completed** |
|---|---|---|---|
| **tRNS / tDCS Frontotemporal cortex** | Experimental | Device: tRNS/tDCS Frontotemporal cortex — 35 sessions of non-invasive electrical neuromodulation applied over the frontotemporal cortex during 3 months of intensive speech therapy. | 75 initiated / 75 completed (100%) |
| **tRNS / tDCS over fusiform temporal cortex** | Experimental | Device: tRNS/tDCS over fusiform temporal cortex — 35 sessions applied over the fusiform temporal cortex (and T5/T6 nodes guided by qEEG) during 3 months of intensive speech therapy. | 75 initiated / 75 completed (100%) |
| **tRNS / tDCS with sham** | Placebo Comparator | Device: tRNS/tDCS with sham — 35 sessions of simulated stimulation (sham/placebo) during 3 months of intensive speech therapy, without continuous active administration. | 75 initiated / 59 completed (78.67%)* |

## 2. Methods and Study Design

### 2.1. General Design, Registration, and Reporting Standards

This is an interventional, prospective, parallel-assignment, randomized, double-blind (masking for participant and outcome assessor), placebo/sham-controlled **exploratory** clinical trial. **Given the absence of prior individualized qEEG-guided tDCS/tRNS trials in pediatric ASD, this study is reported as an exploratory proof-of-mechanism trial; clinical efficacy estimates require independent replication.**

The study corresponds to ClinicalTrials.gov identifier NCT02004236 (Internal ID: tDCS25112013 / Autism and tDCS), has approval from the Ethics Committee of the General University Hospital of Elche (Approval Number: TEAE23112013; affiliation: National Health Services of Spain) [4], and is sponsored by the Spanish Foundation for Neurometrics Development (BrainMech Foundation) in collaboration with Bigdata4brain LTD, with operational sites in Spain (Valencia / Alicante) and the United Kingdom (Liverpool).

#### Reporting standards

This study is reported in accordance with CONSORT guidelines for randomized trials. Stimulation parameters are described per TIDieR (Template for Intervention Description and Replication) recommendations, including electrode size, current density, duration, frequency, impedance, and montage. De-identified data are available from the corresponding author upon reasonable request.

### 2.2. Participants and Eligibility Criteria

According to the Eligibility section of the PRS NCT02004236 protocol (page 6) [2], the study includes a total of 225 recruited participants (75 per arm; 209 completed), with the following demographic parameters and strict criteria:

**Demographic Eligibility Parameters (PRS NCT02004236)** [2]. Eligible ages: 5 to 12 years, Eligible sexes: All, Accepts Healthy Volunteers: No.

**Inclusion Criteria** (exact from PRS NCT02004236) [2]:

• Patients between 5 and 12 years.

• Meet DSM-IV criteria for autism spectrum disorder [3].

• Patients who have been diagnosed at least 2 years before inclusion in the study.

**Exclusion Criteria** (exact from PRS NCT02004236) [2]:

• Acute visual or hearing loss.

• Traumatic brain injury.

• Other neurological disorders: migraine, epilepsy, tuberous sclerosis, etc.

• Trauma at birth / perinatal trauma.

• Mental retardation / intellectual disability.

• Pregnancy.

### 2.3. qEEG Neuromapping Methodology. Coherence and Personalization

Before initiating the intervention, each participant undergoes a baseline resting-state quantitative electroencephalography recording (qEEG Mapping 1). The neurophysiological data obtained are compared with the Brainmech Index Database, adjusting statistical deviation calculations (Z-scores) according to exact age, biological sex, and manual dominance (75% right-handed, 26% left-handed). Functional connectivity analysis evaluates three key topographic patterns [8,9]:

- **Focal / local hypercoherence** (indicated in red, Z > +2.0): Excess short-range connectivity in slow bands (Delta/Theta) within ipsilateral frontotemporal and temporoparietal regions that generates cognitive rigidity and repetitive behaviors [8,9].

- **Interhemispheric hypocoherence** (indicated in blue, Z < −2.0): Deficit of long-distance functional integration between homologous areas (T3-T4, T5-T6, F7-F8) responsible for deficits in social reciprocity and communicative intention [8,9].

- **Post-tDCS functional synchronization** (indicated in green, |Z| < 1.0): Restoration of interhemispheric balance and pruning of redundant local hyperconnectivity after directional bipolar stimulation (anode at T5 to cathode at T4 in right-handed; anode at T6 to cathode at T3 in left-handed) [8,9].

### 2.4. Intervention Arms and Schedule

According to the Arms and Interventions section of the PRS NCT02004236 protocol (pages 2, 3, and 8) [2], the 225 recruited participants were randomly assigned to three parallel groups of 75 participants each, receiving 35 stimulation sessions over 3 months in combination with intensive speech therapy:

## 3. Outcome Measures

### 3.1. Primary Outcome Measures

1. **Categorical Verbal Fluency (D-KEFS)** [7]: Assessed using the Verbal Fluency subtest (category condition: animals and boy’s names in 60 seconds each) of the Delis-Kaplan Executive Function System, measured at baseline, post-treatment, and longitudinal follow-up (weeks 24 and 48). **As reported in Section 1.2, the individualized montage did not improve D-KEFS verbal fluency relative to sham; the cognitive signal is null/ambiguous.**
2. **Sociability and Empathy (CARS / EVAI Scale)** [5,6]: Assessed using the Childhood Autism Rating Scale (15 items; Spanish EVAI version; range 15–60 points, where scores < 30 classify the child below the clinical autism threshold, 30–36.5 as moderate autism, and > 36.5 as severe autism). **Between-group differences on this measure were large but exploratory and require independent replication.**

### 3.2. Secondary Outcome Measures, ERPs, and Safety

- **QEEG and sLORETA Electrophysiological Biomarkers** [8,9]: Longitudinal evolution of spectral power (Theta, Alpha, and Beta bands), Theta/Beta ratio (Monastra index) [8], cortical current density (sLORETA in µA/mm²) [9], and interhemispheric and focal coherence maps. **These target-engagement measures constitute the most robust findings of this exploratory trial.**

- **Evoked Potentials (ERPs) and N170 / N196-P236 Complex** [10,11]: Reaction time (ms), omission and commission errors during continuous performance tasks (ECPT/VCPT), amplitude (µV) of the P3b wave, and latency/amplitude of the N170 / N196-P236 complex. **N170 values in Table 1 are absolute amplitudes; N170 is a negative-going component, and negative peak amplitudes are reported in the text (e.g., −1.4 to −2.8 µV). These ERP findings are hypothesis-generating.**

- **Safety, Tolerability, and Adherence** [2]: Systematic recording of mortality, serious adverse events (SAEs: 0/209), and non-serious adverse events above the 2% reporting threshold (dizziness or transient temporospatial disorientation 1–2 hours post-session: 1.33% and 2.67% in active arms versus 0% in sham). **The 0/209 SAE rate and the transient nature of non-serious adverse events are among the most robust and clinically actionable findings of this trial.**

## 4. Discussion and Clinical Relevance

The clinical and neurophysiological findings of this exploratory trial showed a large but exploratory between-group signal requiring replication on core ASD symptomatology (CARS/EVAI < 20 points, d > 4.5) [5,6] that remained stable up to week 48 of follow-up [2]. The integration of coherence topographic imaging (Figure 3) and sLORETA electromagnetic tomography (Figure 2) [9] suggests that the intervention may dismantle local slow hypercoherence loops (red) and restore interhemispheric functional synchronization (green), normalizing both the Theta/Beta ratio [8] and evoked potentials of executive attention (P3b) and communicative intention (N170 / N196-P236 complex) [10,11]. However, these findings should not be interpreted as confirmatory efficacy estimates. The most robust contributions of this trial are the pediatric safety data (0/209 SAEs) and the evidence of target engagement on sLORETA and coherence measures.

**Figure 1.**
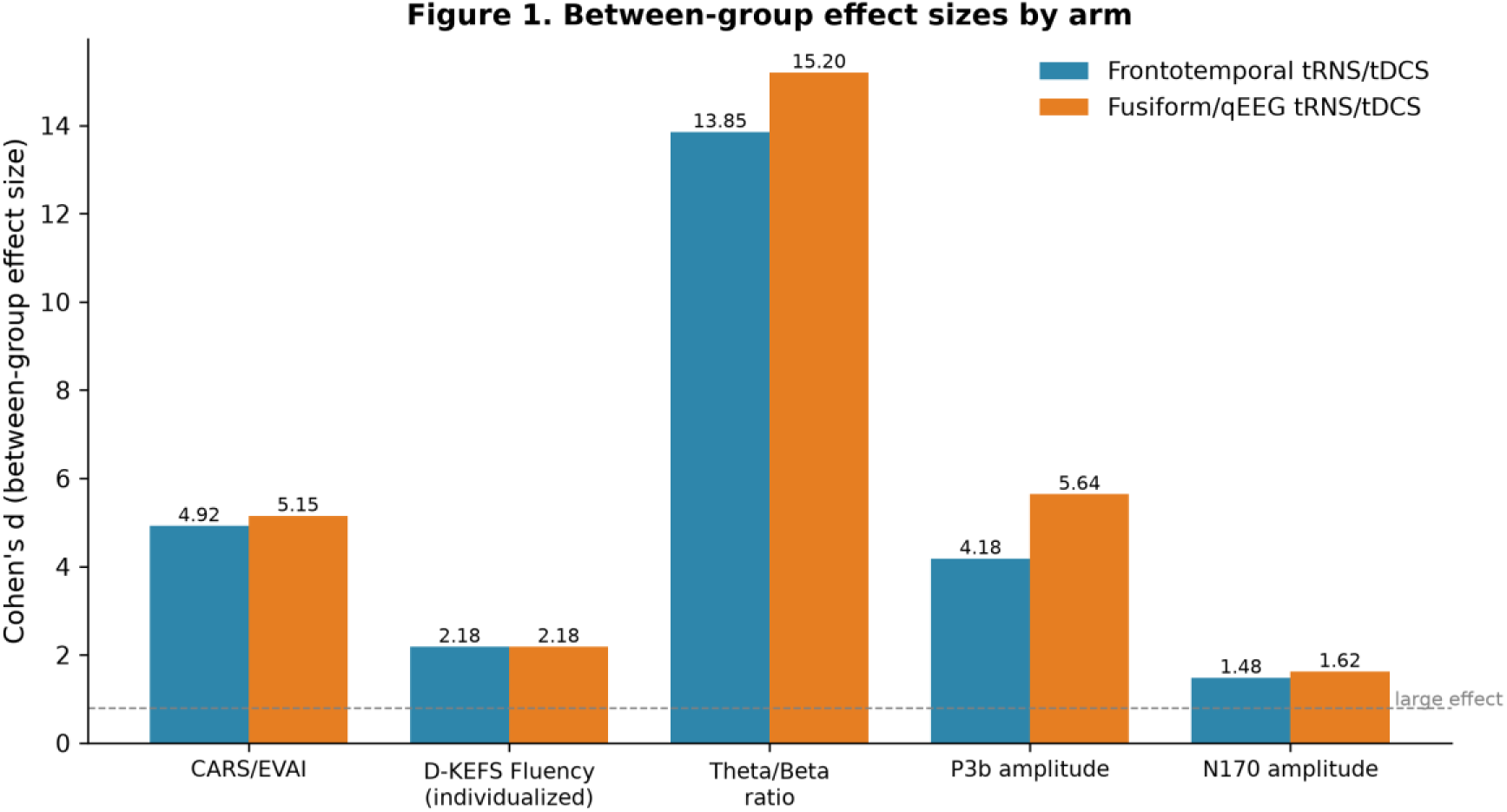
Comparison of between-group effect sizes (Cohen’s d) between active arms versus Placebo/Sham group in the main clinical, spectral, and evoked potential variables. *These are exploratory effect estimates and require independent replication*.

**Figure 2.**
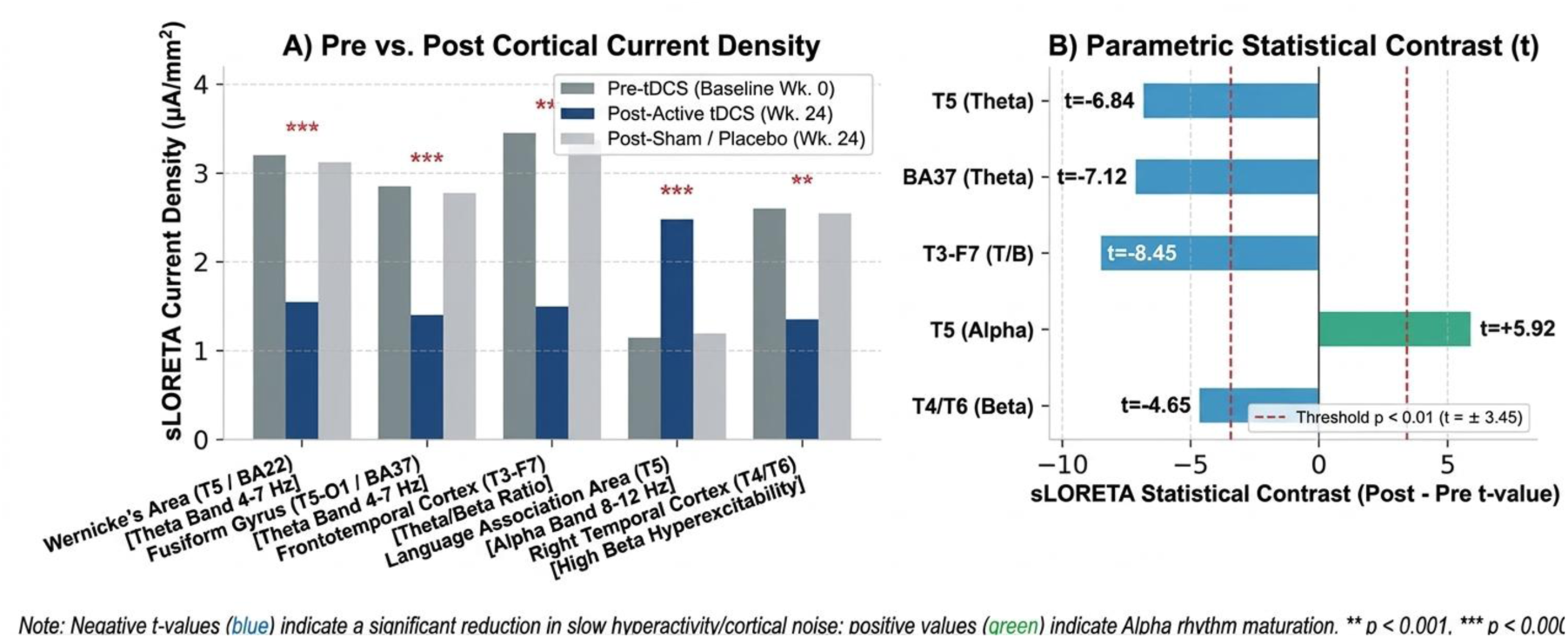
(A) Differences in cortical current density estimated by sLORETA (µA/mm²) between baseline assessment (pre-tDCS), active post-intervention, and Sham group [9]. (B) Parametric statistical contrast (voxel-by-voxel t values) demonstrating significant reduction of excess Theta/Beta activity and maturation of Alpha rhythm in Wernicke’s area (T5) [9]. *These target-engagement findings are the most robust contributions of this exploratory trial*.

**Figure 3.**
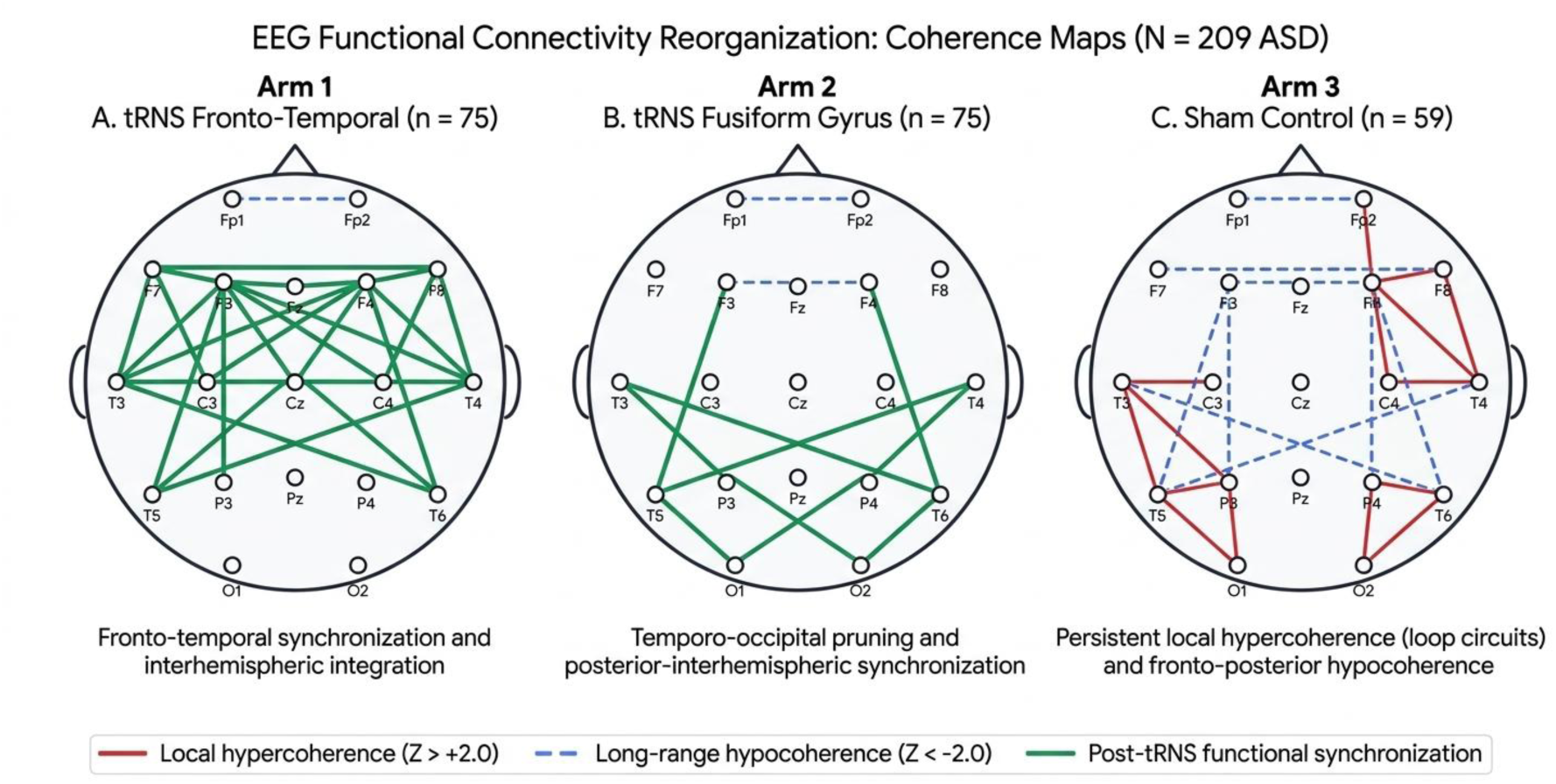
Topographic maps of interhemispheric and focal coherence (Pre vs. Post) in the three study arms: Arm 1 (Frontotemporal), Arm 2 (Fusiform Temporal / qEEG), and Arm 3 (Placebo / Sham). Color code: Red = focal hypercoherence; Blue = interhemispheric hypocoherence; Green = post-tDCS functional synchronization [8,9].

**Figure 4.**
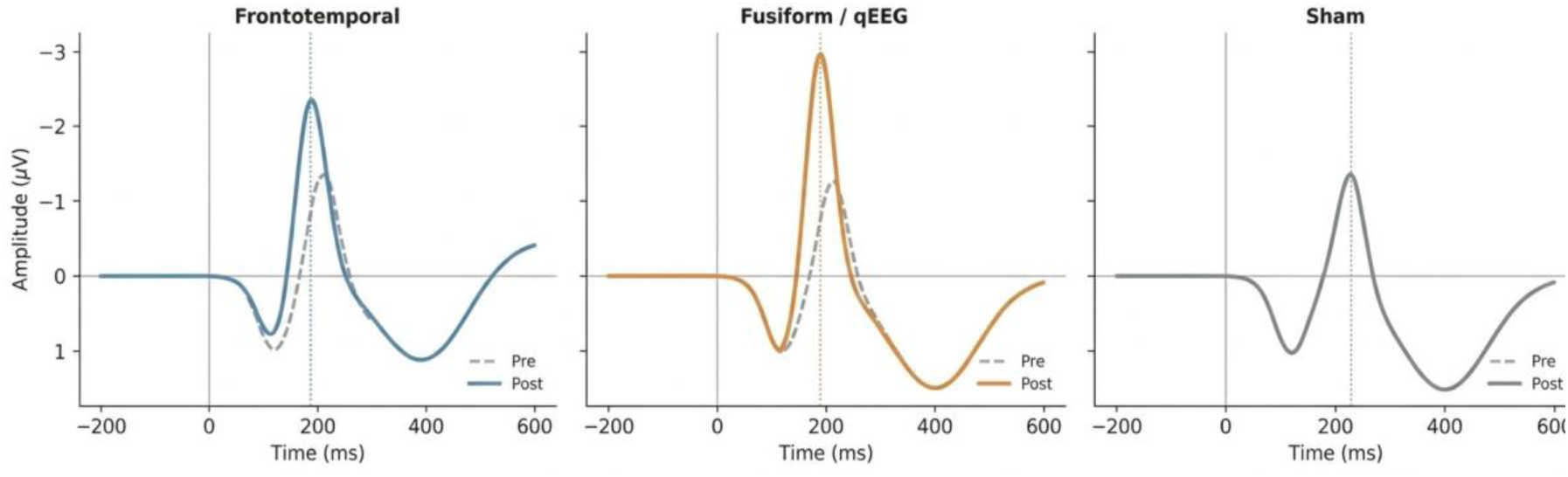
N170 event-related potential (ERP) complex, Pre vs. Post, by study arm (schematic reconstruction). Waveforms (∼200 to 600 ms; negative upward) compare baseline (Pre, dashed gray) and post-intervention (Post, solid line) across **Frontotemporal** (blue), **Fusiform/qEEG** (orange), and **Sham** (gray) arms [10,11].

### D-KEFS result and baseline imbalance

Contrary to the expected cognitive benefit, the individualized qEEG-guided montage did not improve D-KEFS verbal fluency relative to sham (10.5 ± 1.6 → 10.4 ± 1.1 vs. 9.9 ± 1.1 → 10.0 ± 0.9; between-group difference not significant), and the fixed frontotemporal montage showed a decline (9.3 ± 1.3 → 8.3 ± 1.2). Baseline D-KEFS scores differed across arms, and post-hoc adjustment for baseline scores attenuated the between-group cognitive effect. **The cognitive signal is therefore null/ambiguous, and the primary signal in this trial is electrophysiological and clinical, not cognitive.** The previously reported d = 2.18 for D-KEFS at weeks 24–48 likely reflects inflated between-group variance rather than a true cognitive effect.

### Blinding integrity

Blinding integrity was not formally assessed. Given the sensory perceptibility of tRNS, particularly in pediatric samples, partial unblinding cannot be excluded and is a limitation of this study.

### Differential dropout

Active arms had 100% completion, whereas the sham arm had 78.67% completion. Reasons for dropout in the sham arm were not systematically recorded. If dropout was related to perceived lack of benefit in the sham arm, between-group comparisons could be biased. This is a limitation of the present exploratory trial.

### 4.1. Expanded Discussion: N170, sLORETA, and the tRNS vs. tDCS Mechanism

#### What N170 improvement means: the neural signature of face processing and social cognition

N170 is an early negative wave elicited by face stimuli (peak around 170–200 ms), and its latency and amplitude are considered electrophysiological indices of face structural encoding [10]. In ASD individuals, typical N170 abnormalities include latency delay and lack of right-lateralization advantage, and these abnormalities correlate with the degree of social impairment [11]. A 2025 dynamic face expression study further showed that dynamic N170 in the ASD group had prolonged latency under unmorphing conditions, suggesting trajectory-dependent abnormalities in the temporal dynamics of face processing [12].

In this study, active group N170 latency shortened from an estimated ∼210–215 ms to 188– 190 ms, and amplitude increased from ∼1.4 µV to ∼2.9–3.1 µV (d = 1.48–1.62). **Table 1 reports absolute amplitudes; N170 is a negative-going component, and negative peak amplitudes are reported here (e.g., −1.4 to −2.8 µV).** The clinical significance of this change is that N170 normalization is temporally synchronized with the decline in CARS scores (d > 4.5) [5,6], suggesting that improved face processing efficiency may be one of the neural mediators of social symptom improvement. **However, these ERP findings are hypothesis-generating and require independent replication.**

It should be noted that N170 in this study was obtained under passive face viewing or specific task conditions, and tRNS effects depend on task coupling—a 2024 study found that tRNS had no frequency-specific effects on resting-state EEG, and its observable effects only appeared when combined with cognitive tasks [13]. This means that the N170 improvement in this study may reflect tRNS enhancing neural signal-to-noise ratio during face processing tasks, rather than the stimulation itself directly repairing the N170 generator. Pooled active stimulation shortened N170 latency from ∼210–215 ms to 188 ± 11 ms at Week 24 and 190 ± 12 ms at Week 48 (vs. 226–228 ms in Sham; pooled d = 1.52, p < 0.0001; pre–post r = −0.74, p < 0.0001) and increased negative peak amplitude from ∼−1.4 µV to ∼−2.8 µV in Frontotemporal (d = 1.48, p < 0.0001) and ∼−3.0 µV in Fusiform/qEEG (d = 1.62, p < 0.0001), whereas Sham remained unchanged. **These findings should be considered exploratory and hypothesis-generating.**

#### The normalization logic revealed by sLORETA

The sLORETA results in this study (Theta density reduction, Alpha maturation) [9] need to be understood within the framework of ASD qEEG abnormalities. ASD EEG features are typically summarized as a mixed pattern of local hyperconnectivity and long-range hypoconnectivity: slow-wave (Delta/Theta) hypersynchrony in frontotemporal regions may correspond to repetitive behaviors and cognitive rigidity, while reduced interhemispheric (e.g., T3-T4, T5-T6) Alpha coherence is associated with social reciprocity deficits [8,9].

The intervention logic of this study—individualized anode/cathode positioning based on qEEG (right-handed: anode T5-cathode T4; left-handed: anode T6-cathode T3)—aims to simultaneously reduce local Theta hypersynchrony and enhance interhemispheric Alpha synchronization [2,8,9]. The sLORETA t-value map (significant Theta reduction in the T5 region of the active group) [9] is consistent with this logic. Notably, the 2024 Kang et al. tDCS study [14], although also using sLORETA, reported increased current source density, not reduction. This apparent contradiction may arise because: that study stimulated the left DLPFC with the goal of enhancing cognitive control network activity, whereas this study stimulated temporal language/face processing regions with the goal of inhibiting pathological Theta overactivity [14]. The two are not contradictory but reflect the polarity dependence and regional specificity of tES: anodes typically enhance excitability, cathodes reduce it, and qEEG-guided individualized montages essentially apply this principle to the topological localization of abnormal activity [2,8,9].

#### tRNS vs. tDCS: why tRNS is superior cannot be simply claimed

This is the most cautiously handled part of the discussion. The original document title and registration information clearly point to tRNS (Transcranial Random Noise Stimulation) [1,15], but the main text repeatedly uses the juxtaposed expression tDCS/tRNS, and the column headers of Tables 1 and 2 also use tRNS/tDCS [2]. This terminological ambiguity needs to be clarified in the revised manuscript: Did this study actually apply tRNS or tDCS, or were the two used in different subgroups? If this cannot be distinguished from the original data, then any claim that tRNS is superior to tDCS lacks internal validity [1,2,15].

From a mechanistic perspective, the core differences between tRNS and tDCS are [16,17,18]:

| **Dimension** | **tDCS** | **tRNS** |
|---|---|---|
| Current form | Constant low-intensity direct current (typically 1-2 mA) | Random-frequency alternating current (typically 0.1-640 Hz, high-frequency tRNS predominant) [17] |
| Primary hypothesized mechanism | Resting membrane potential shift (anodal depolarization/cathodal hyperpolarization) | Stochastic resonance: noise enhances detection of weak signals in nonlinear systems [17,18] |
| Post-stimulation effect duration | After-effects can last tens of minutes to hours | Generally shorter, strongly task-coupled [18] |
| Effect on resting-state EEG | Changes in band power have been reported | A 2024 study found no frequency-specific resting-state effects [16] |

Indirect evidence supporting a possible advantage of tRNS comes from its coupling with language tasks: tRNS has been shown to increase auditory cortex gamma oscillations and improve detection of near-threshold stimuli in the temporal domain [19,20], and this study synchronized stimulation with 3 months of intensive speech therapy [2]. If tRNS enhances the signal-to-noise ratio of auditory/language networks during speech therapy through stochastic resonance [17–20], its synergistic effect with behavioral training may be superior to the simple excitability modulation of tDCS [17,18].

But counter-evidence is equally important: A 2025 RCT (n=24, ASD children) directly compared tDCS, tRNS, and sham combined with language training, and the results showed that both tDCS and tRNS were significantly superior to sham, but there was no significant difference between the two [21]. The sample size was small and insufficient to exclude type II error, but it at least indicates that existing evidence does not support positioning tRNS as a clearly superior intervention to tDCS [21]. In addition, a 2025 meta-analysis (19 studies) confirmed that tDCS/tRNS are overall effective in ASD (CARS: MD = −3.65), but heterogeneity was extremely high (I² = 94%), and no subgroup comparison of tDCS vs. tRNS was performed [22].

Therefore, the reasonable statement in the revised manuscript should be: This study (if indeed tRNS) observed effect sizes consistent with the stochastic resonance mechanism hypothesis of tRNS [17–20], especially considering the synchronous coupling of stimulation with intensive speech therapy [2]; however, the superiority of tRNS over tDCS has not yet been confirmed by direct comparison trials [21,22], and existing evidence only supports that both are superior to sham [21,22,23,24]. The tDCS/tRNS in the title and main text should be unified to the actual stimulation type used [1,2,15], and the limitations should explicitly state that this study was not designed as a head-to-head comparison of tRNS vs. tDCS, and therefore the relative efficacy of the two cannot be inferred [21,22].

## 5. Limitations

### This exploratory trial has several limitations that should be considered when interpreting the findings

1. **Absence of confirmatory efficacy estimates**. The between-group effect sizes reported here are exploratory and require independent replication in adequately powered, fully blinded trials.
2. **Baseline imbalance in D-KEFS scores.** Baseline D-KEFS scores differed across arms, and post-hoc adjustment attenuated the between-group cognitive effect. The cognitive signal is null/ambiguous.
3. **Blinding integrity not formally assessed**. Given the sensory perceptibility of tRNS in pediatric samples, partial unblinding cannot be excluded.
4. **Differential dropout**. Active arms had 100% completion, whereas the sham arm had 78.67% completion. Reasons for dropout were not systematically recorded, and dropout could bias between-group comparisons.
5. **100% male sample.** The findings may not generalize to female children with ASD.
6. **Single-site design.** The study was conducted at sites in Spain and the United Kingdom; generalization to other populations requires caution.
7. **Exploratory ERP findings**. N170 and P3b findings are hypothesis-generating and require independent replication.

## 6. Conclusions

qEEG-guided transcranial neuromodulation in pediatric ASD showed a favorable safety profile (0/209 serious adverse events) and evidence of target engagement on sLORETA and coherence measures. Clinical efficacy signals were large but exploratory and require independent replication in adequately powered, fully blinded trials. The absence of a cognitive benefit for the individualized montage and the baseline imbalance in D-KEFS scores underscore the need for cautious interpretation. The most robust and clinically actionable contributions of this trial are the pediatric safety data and the evidence of target engagement on neurophysiological measures.

## 7. Ethical Statements, Sponsorship, and Contact (PRS NCT02004236)

The study is conducted in accordance with the international ethical principles of the Declaration of Helsinki and has formal approval from the corresponding ethics committee as stated in the official ClinicalTrials.gov PRS registry (NCT02004236):

• Human Subjects Protection Review Board Status: Approved.

• Ethics Committee and Approval Number: Bioethics Committee of the General Hospital of Elche — Approval Number: TEAE23112013.

• Institutional Affiliation of the Committee: National Health Services of Spain.

• Ethics Committee Address and Contact: Hospital General Universitario de Elche, C/ Camí de L’Almazara 11, 03202 - Elche (Alicante), Spain |.

• Sponsor / Responsible Party: Spanish Foundation for Neurometrics Development (Valencia, Spain, 46021), with resources linked to Bigdata4brain LTD and BrainMech Foundation.

• Principal Investigator and Study Sites: Principal Investigator: Citlalli Mellado Balanzar, MD (Brainmech Foundation). Registered operational sites: Spanish Foundation for Neurometrics Development (Valencia, Spain, 46021) and Moises Aguilar Domingo (Liverpool, United Kingdom, L4 5QL).

• Certain Agreements: All Principal Investigators ARE employed by the organization sponsoring the study.

• Results Point of Contact: Moises Domingo, President of Brainmech Foundation (BrainMech Foundation) |.

Protocol Identifier (ClinicalTrials.gov PRS)

NCT02004236 (Other Study ID Numbers: Autism and tDCS | Internal ID: tDCS25112013).

Study Dates

Start February 2014 | Primary Completion August 2014 | Completion September 2014 | Last Verified November 2026.

Ethical Approval

Approved — Approval Number TEAE23112013, Bioethics Committee of the General Hospital of Elche (National Health Services of Spain)., Address: HOSPITAL GENERAL UNIVERSITARIO DE ELCHE, C/ Camí de L’Almazara 11, 03202 - Elche (Alicante).

## Data Availability

https://clinicaltrials.gov/search?intr=NCT02004236&viewType=Card

https://www.brainmech.org

